# Machine Learning-Based Pattern Recognition of Risk Factors for Low Back Pain among Adolescent Cricket Players in Dhaka City

**DOI:** 10.1101/2025.05.06.25327129

**Authors:** Marzana Afrooj Ria, Tasrima Trisha Ratna, Shudeshna Chakraborttye Purba, Rubal Kar, Mohoshina Karim, Md Osman Ali, Erfat Jaren Chaity, Shahadath Hossen, Joynal Abedin Imran, Shahriar Hasan

## Abstract

Low back pain (LBP) is common among adolescent cricketers, often due to repetitive lumbar stress. This study investigated LBP among 450 adolescent cricketers in Dhaka City in Bangladesh, considering a range of factors, including sociodemographic characteristics, game-related activities, preventive practices, and LBP-related history. Several machine learning (ML) algorithms were applied to classify LBP severity, including K-Nearest Neighbors, Random Forest, Logistic Regression, and Support Vector Machine (SVM). The severity of LBP was categorized into three classes such as no pain, mild pain, and moderate pain owing to insufficient data in the severe pain category. Among the tested models, SVM with a sigmoid kernel demonstrated the best performance, achieving the highest performance metrics of test accuracy (87.6%), precision (90%), recall (87.6%), and F1-score (87.1%). In addition, regression analysis was performed to identify the contributing factors associated with LBP. Significant statistical correlations were evident between LBP and variables such as gender, educational background, family income, duration of practice, warm-up and cool-down protocol, and past history of LBP. These findings emphasize the importance of early preventive measures in lessening LBP risk among young cricket players. Overall, this study demonstrates the utility of ML and regression models in identifying sports injury risk patterns, supporting data-driven prevention and management strategies, and providing a baseline for future research in this field.

## Introduction

Low back pain (LBP) in adolescent athletes, particularly in cricketers, is growing concern due to repetitive spinal stress. Adolescents are highly susceptible since due to their developmental stage and sports participation [1]. Repeated spinal movements in games such as cricket are responsible for LBP in this case, with a rate of up to 30% of adolescents affected [2]. Female adolescents and children have a greater incidence of LBP compared to their male counterparts [3]. Despite its prevalence, effective LBP prevention is limited. The complex nature of LBP, stemming from both biomechanical and psychosocial etiologies, hinders detection and intervention [4].

In cricket, there is severe mechanical stress on the lumbar spine through repeated hyperextension, rotation, and lateral flexion, significantly raising the risk of LBP [5,6]. Immature spines in junior bowlers are especially susceptible [6]. Repeated acts of bowling have resulted in structural spine injuries such as spondylolysis through cyclic extension loading [7], and therefore cricket is also a high-risk sport for LBP. This highlights the urgent need for injury prevention and biomechanical monitoring. LBP in young athletes can limit physical functioning, hinder return to sport, and have a detrimental effect on emotional well-being and identity. Adolescents often normalize pain, leading to underreporting and undertreatment, with detrimental effects. LBP also strains peer and coach relationships [2]. Recurrent LBP in development years can contribute to adulthood risk of chronic pain and sick leave from school [1].

Thus, early detection of young athletes with potential LBP risk factors is essential to guarantee effective prevention and interventions, helping to preserve both sports competence and future health [1]. It also allows coaches and medical professionals to establish personalized training and rehabilitation programs to reduce injury [4]. Isokinetic testing and electromyography are good screening instruments in identifying at-risk individuals [4]. Interventions made in a timely manner can avoid long-term consequences such as persistent pain and impaired performance [2].

Furthermore, traditional manual protocols for detecting injury-risk factors in athletes have limited capacity and cannot identify complex patterns [8]. In contrast, machine learning (ML) offers a data-driven approach to identify subtle risk factors by processing large datasets, including information on training history, biomechanics, and psychological factors, which are difficult to gather manually [9]. This methodology is particularly suitable for contexts like adolescent cricket players in Dhaka city, where studies on LBP-associated factors are scarce [6]. The cricket biomechanics in general and fast bowling in particular account for increased LBP risk [10]. Risk factors are stratified through the use of ML models so prediction of the probability of LBP is possible, and health professionals can forecast risks and individualize prevention and rehabilitation strategies [8,9].

There are no known predictive models with ML to be used in screening LBP in adolescent cricketers in Dhaka. The aim of this study is to fill this gap with ML and regression analysis to define important risk factors in the setting of Bangladesh. The findings will inform health professionals, coaches, and sports managers about optimal prevention and management strategies for LBP in young cricketers. This study can also serve as a model for future research on sports injuries using ML applications in other regions or sports disciplines.

## Materials & Methods

### Study population

For this study, 450 adolescent cricket players data were collected using purposive sampling technique across various cricket clubs (BKSP, Lt. Sheikh Jamal cricket Academy, Kola Bagan Krira chakra, Khelaghar Cricket Academy and City club) in Dhaka City, with the primary focus on identifying the risk factors associated with the development of LBP. The data collection was conducted from 25 February 2024 to 28 November 2024.

### Ethical consideration

This study adhered to the ethical principles of the Declaration of Helsinki. This study was also approved by the IRB of the Institutional Review Board (IRB) of the National Institute of Traumatology and Orthopaedic Rehabilitation (NITOR/PT/93/lRB/2024/05). Participants were fully informed of the study’s purpose, methodology, and procedures. The participants had the right to withdraw from the interview completely at any time. Confidentiality and anonymity were ensured. No physical specimens were collected.

### Outcome variables

To facilitate the data collection and classification process, LBP severity was categorized into three distinct classes, such as Class 0: No pain (No LBP), Class 1: Mild pain, Class 2: Moderate pain. Specifically, 150 participants were categorized as Class 0 (No pain), while the remaining 300 participants were categorized as Class 1 (Mild pain) and Class 2 (Moderate pain).

### Explanatory variables

The dataset included a variety of features capturing the demographic, physical, and training-related characteristics of adolescent cricket players. Data were collected through structured surveys and physical assessments, and were organized into the following categories:

- Sociodemographic factors: Age, gender, education, monthly family income, and body mass index.
- Games-related factors: Playing position, playing experience, and duration of practice per week.
- Preventive measures factors: Warm-up before sports activity, duration of warm-up, cooldown after sports activity, and duration of cool down.

Players’ ages and heights ranged from 11 to 19 years and from 120 to 180.3 cm, respectively. This information provides insight into their physical stature, which may influence biomechanics during cricketing activities. Similarly, players’ weight can significantly affect the strain on the musculoskeletal system during high-intensity activities like cricket. In this study, players’ weights ranged from 20 to 80 kilograms.

The players’ Body Mass Index (BMI) varied between 8.53 and 29.81. This measure was split into three categories: underweight (BMI < 18.5), normal (BMI 18.5 - 24.9), and overweight (BMI ≥ 25), with implications for the overall risk of injury to the player. The educational level was divided into four categories: J.S.C./8th grade, S.S.C./O-levels, H.S.C./A-levels, and undergraduate. This variable is important as education may reflect underlying socio-economic factors influencing players’ overall health and physical activity.

Position of play was another factor of significance, following categories such as right-hand batsman, left-hand batsman, spin hand bowler, pace hand bowler, wicket-keeper, and all-rounder. These roles dictate the type of biomechanical stress players experience; bowlers particularly pace bowlers, face a higher LBP risk due to repetitive movements. The players were classified in terms of their playing experience among three categories, 1-3 years, 4-6 years, and 7-10 years. The players’ practice duration per week (hour) was classified in three categories viz. <10 hours, 10-20 hours, and >20 hours.

Training habits are captured through the doing and duration of warm-up and cooldown. Warm-up before and cooldown after the sports activity were categorized into ‘always’, ‘often’ and ‘sometimes’. The warm-up duration ranges from <10 minutes to >15 minutes, while cooldown time is similarly categorized into <10 minutes, 10-15 minutes, and >15 minutes. These factors are critical as appropriate warm-up and cooldown routines are known to reduce the risk of injury and improve muscle recovery. Lastly, past history of LBP is documented either “Present” or “Absent”.

In summary, the machine learning model for LBP severity classification was developed using an integrated set of attributes, including sociodemographic factors, game-related factors, preventive, and LBP related factors. The model’s performance was evaluated using 10-fold cross-validation.

### Selection of Algorithms

The most popular machine learning-based classification models include Logistic Regression (LR), K-Nearest Neighbors (KNN), Random Forest (RF), and Support Vector Machine (SVM), whereas deep learning-based classification models are Convolutional Neural Networks (CNN) and Deep Neural Networks (DNN). These machine learning-based model were chosen in this study because of their effectiveness in handling structured data in contexts with a small number of samples. The deep learning models are in need of a large volume of data to automatically extract and learn intricate feature representation efficiently [11].

This study evaluates the performance of four machine learning classifiers: SVM, RF, KNN, and LR for the classification of LBP severity among adolescent cricket players. The dataset was balanced, consisting of 150 instances in each of the three classes: No pain, Mild pain, and Moderate pain. Of the above-mentioned machine learning algorithms, KNN is a non-parametric model whereas RF and SVM have good classification properties and are suitable for non-linear decision boundary modelling.

### Logistic Regression

LR is a widely used statistical method for binary and multiclass classification in medical research [12]. It models the probability of class membership using the logistic function, which is a type of sigmoid function. This function assumes a linear relationship between the independent variables and the log-odds of the outcome. It is efficient, interpretable, and works best when the data has a linear decision boundary. It is sensitive to multicollinearity and may underperform in complex or nonlinear datasets unless extended with interaction terms or regularization. In this study, logistic regression served as a baseline model. While its simplicity allowed clear insights into individual predictors of low back pain, it was less effective in capturing nonlinear associations compared to tree-based or kernel-based models.

### K-Nearest Neighbours

KNN is a non-parametric, instance-based learning algorithm commonly used in both classification and regression tasks, particularly within structured medical data contexts due to its simplicity and effectiveness in handling health-related patterns [13]. It predicts outcomes by identifying the *k* closest data points to a given instance based on a distance metric, typically Euclidean distance:

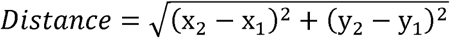

KNN performs well on small to moderately sized datasets with low to medium dimensionality and balanced class distributions. It is particularly useful when the data exhibits local patterns or clustering. However, its performance can degrade in high-dimensional or noisy datasets, and it becomes computationally expensive with large datasets due to its lack of a training phase.

In this study, KNN was applied during the model evaluation phase to explore pattern similarity among adolescent cricket players. Although it offered interpretable baseline results, it was less effective than more robust classifiers like SVM in capturing complex relationships between low back pain and its contributing factors.

### Random Forest

Random Forest (RF) is an ensemble learning algorithm [14] that builds multiple decision trees and combines their outputs to improve classification accuracy and reduce overfitting. Each tree is trained on a random subset of the data and features, and final predictions are made by majority voting for classification or averaging for regression. It is well-suited for handling datasets with nonlinear relationships, mixed data types, and moderate levels of noise. It provides good performance even without extensive parameter tuning and offers feature importance scores, aiding interpretability. However, its complexity can increase with many trees, leading to slower predictions and less transparency compared to simpler models.

In this study, Random Forest demonstrated strong classification performance in identifying low back pain patterns, particularly due to its robustness against overfitting and ability to model interactions between risk factors.

### Support Vector Machine (SVM)

SVM is a supervised learning machine learning algorithm with excellent performance in multiclass classification even when it has been trained on relatively small data sets. It is a human-interpretable model and has good generalizing capacity on small data samples and is less susceptible to overfitting in comparison to ensembled-based models such as RF [15,16,17]. SVM generates optimal separating decision boundaries, called hyperplanes, in a high-dimensional feature space to classify data into their respective classes. These hyperplanes are maximally positioned between classes. In other words, the hyperplanes are put in such a way that they are maximally distant from data points of any class. These data points are called support vectors. Maximizing this distance between classes is what aims to help improve the model’s performance in generalizing new data [15].

Several mathematical functions known as kernels are employed to deal with non-linearly separable data like linear kernel, polynomial kernel, radial basis function (RBF), and sigmoid. These kernels enable the mapping of original data to a higher-dimensional feature space, where it becomes linearly separable. Mathematical representation of these kernels is provided in Table 1 and a representation of SVM-based decision boundaries for these kernels is shown in Fig.1.

**Fig. 1.**
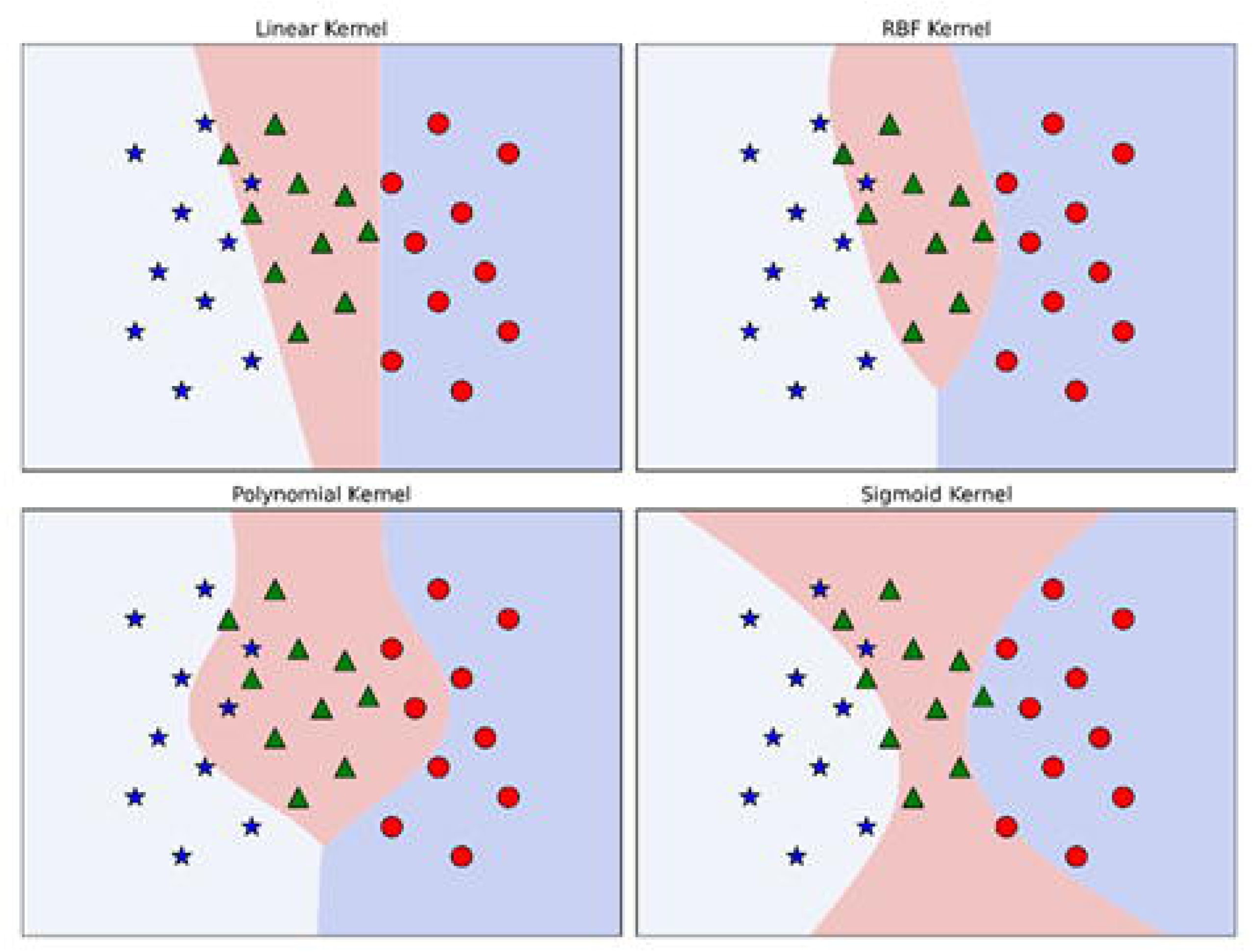
An illustration of SVM decision boundaries using different kernel functions.

**Table 1.**
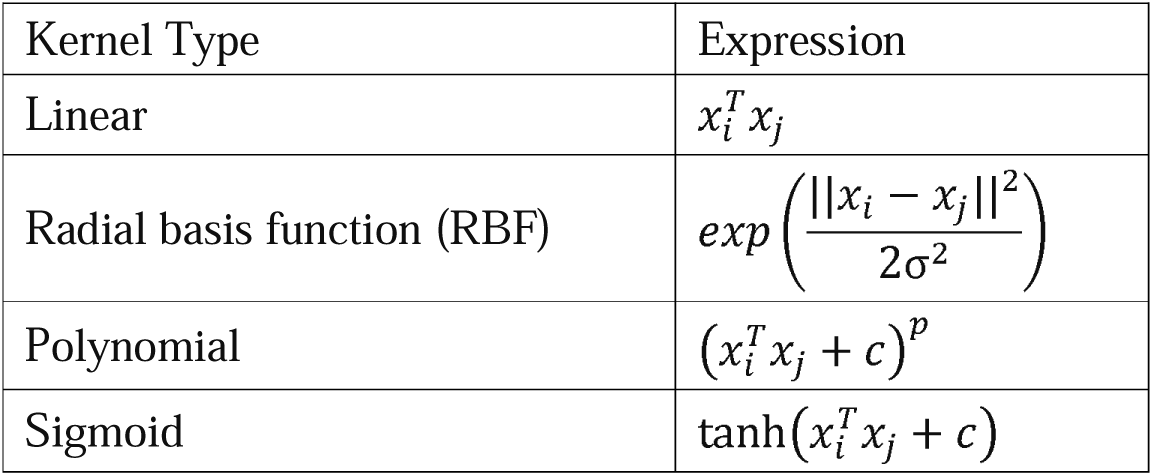
SVM kernel functions and their mathematical expressions.

In this work, several approaches were undertaken to minimize overfitting and improve generalization. For instance, ten-fold cross-validation was applied to ensure balanced evaluation across the dataset. The dataset itself was balanced with equal class representation to avoid bias. Additionally, hyperparameters for each model were optimized using grid search to identify the most effective configurations. These methods collectively helped reduce variance and ensure more reliable classification performance.

### Performance Metrics

To ensure reliable model evaluation given the limited dataset size, ten-fold cross-validation (CV-10) was used. The classifier’s effectiveness was assessed using commonly used evaluation metrics, including accuracy, precision, recall, and F1-score, which are also necessary to comprehensively analyze performance in the presence of class imbalance. These metrics are calculated based on the number of true positive (TP), true negative (TN), false positive (FP), and false negative (FN) predictions, as defined below:

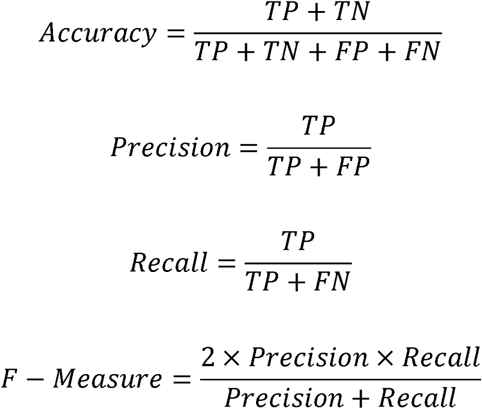

All performance metrics were computed as averages across the folds, providing stable estimates of each model’s generalization performance.

## Result

The research result was analyzed by two different types of analytical method; among them one was ML-based classification models which included KNN, RF, LR, and SVM model. Another analytical model was binary logistic regression model.

Among the classifiers, SVM achieved the highest overall performance. After extensive grid search-based hyperparameter tuning, the best configuration was found with a sigmoid kernel, C = 1, and gamma = 0.01. As shown in Table 2, SVM attained a test accuracy of 87.6% and a macro-averaged F1 score of 87.1%, outperforming the other models. RF and LR followed closely, with F1 scores of 85.4% and 85.9%, and test accuracies of 85.6% and 86.0%, respectively. KNN, although achieving 100% training accuracy, had the lowest generalization ability, with a test accuracy of 80.4% and an F1 score of 78.9%, indicating overfitting.

**Table 2:** Overall Algorithm Performance Comparison.

| Algorithm | Train Acc | Test Acc | Precision | Recall | F1 Score |
| --- | --- | --- | --- | --- | --- |
| KNN | **1.000** | 0.804 | 0.837 | 0.804 | 0.789 |
| RF | 0.967 | 0.856 | 0.863 | 0.856 | 0.854 |
| LR | 0.898 | 0.860 | 0.866 | 0.860 | 0.859 |
| **SVM** | 0.884 | **0.876** | **0.900** | **0.876** | **0.871** |

To further asses the performance of the SVM classifier in detail, class-wise results were evaluated and are summarized in Table 3. Class 0 (No pain) achieved perfect classification, with 100% precision, recall, and F1-score. Class 1 (Mild pain) was also well classified, with an F1 score of 84.0%. In contrast, Class 2 (Moderate pain) exhibited more classification difficulty, with a precision of 95.5% but a lower recall of 66.0%, resulting in an F1 score of 77.4%. These results indicate that while the SVM model performs well overall, it faces significant challenges in accurately identifying moderate pain cases (Class 2).

**Table 3:** Class-wise Performance Metrics.

| Class | Train Acc | Test Acc. | Precision | Recall | F1 Score |
| --- | --- | --- | --- | --- | --- |
| 0 | 1.000 | 1.000 | 1.000 | 1.000 | 1.000 |
| 1 | 0.974 | 0.967 | 0.745 | 0.967 | 0.840 |
| 2 | 0.679 | 0.660 | 0.955 | 0.660 | 0.774 |

Visual performance metrics are shown in Figure 2. In Figure 2(a), training and test accuracies across all folds remain closely aligned, with training accuracy around 88–89% and test accuracy between 81–88%, confirming strong generalization and minimal overfitting. Figure 2(b) displays class-wise accuracy across folds. Class 0 achieved perfect accuracy in all folds, while Class 1 remained stable. Class 2 varied more significantly, with fold-wise accuracy ranging from 45% to 80%, underscoring its classification complexity.

**Figure 2.**
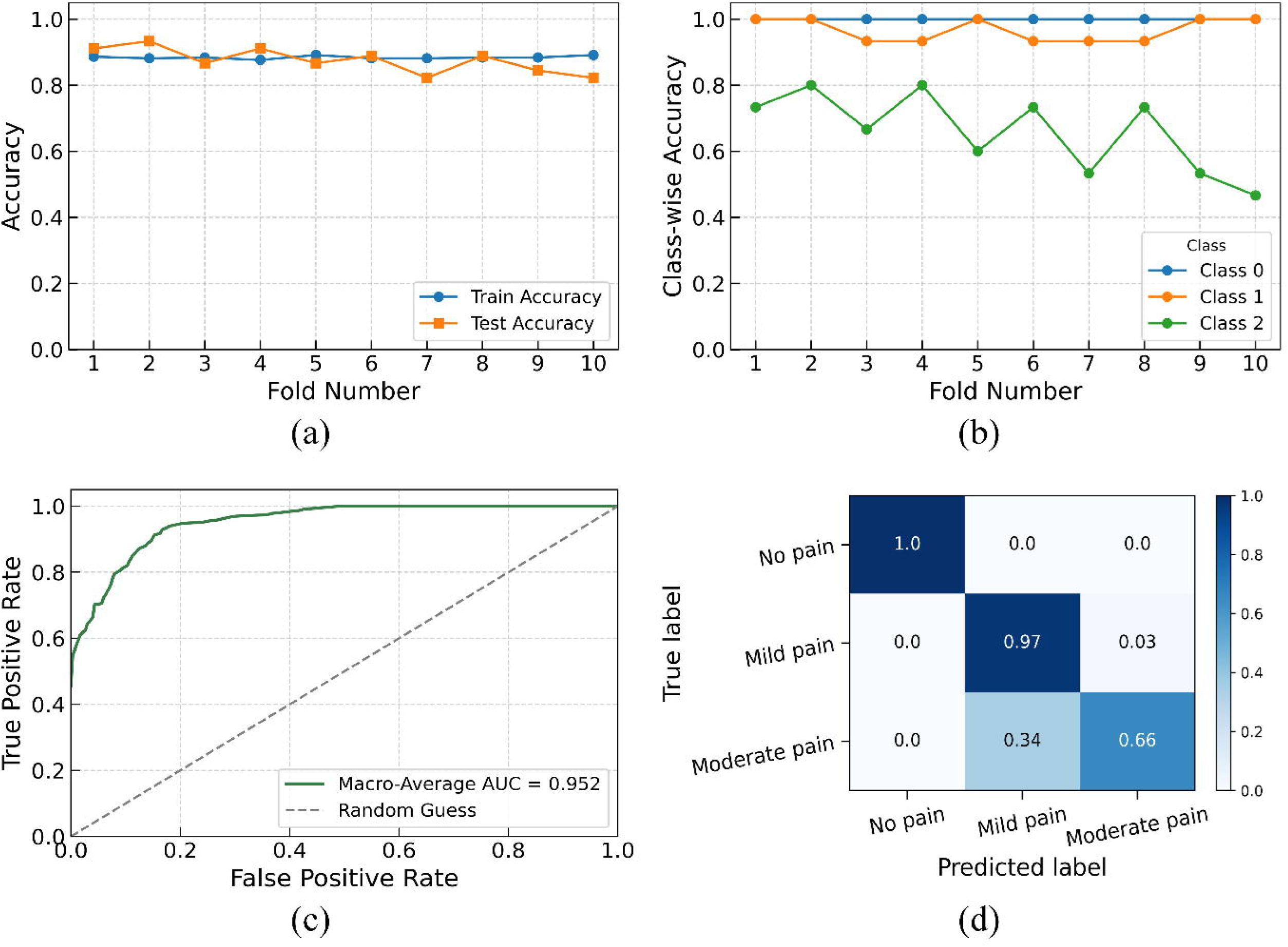
Performance of the SVM classifier with sigmoid kernel (C=1, gamma=0.01) for LBP classification: (a) train and test accuracy across folds, (b) class-wise test accuracy, (c) macro-average ROC curve (AUC = 0.952), and (d) normalized confusion matrix.

Figure 2(c) presents the macro-averaged ROC curve, where SVM achieved an AUC of 0.952, reflecting excellent discrimination among all three classes. The normalized confusion matrix in Figure 2(d) shows 100% correct classification for Class 0, 97% for Class 1, and 66% for Class 2. Misclassification of 34% of Class 2 samples as mild pain illustrates the overlap in features between moderate and mild pain cases and signals a need for enhanced feature differentiation.

### Sociodemographic, games and preventive measures related characteristics of the participants

This study revealed that several sociodemographic characteristics are significantly associated with the prevalence of low back pain (LBP). A strong association exists for both age (p<0.001) and educational level (p<0.001), showing that LBP becomes more common as players get older and advance in their schooling. Monthly family income is also a significant factor (p<0.001), where participants from the lowest income group reported substantially less LBP than those from higher-income families. Body Mass Index (BMI) showed a significant association (p=0.011).

**Table 4.** Distribution of sociodemographic characteristics of the participants (n=450)

| Characteristics | Low back pain |  | Chi-square | P-value |
| --- | --- | --- | --- | --- |
|  | Present (%)<br>(n = 300) | Absent (%)<br>(n = 150) |  |  |
| Age |  |  |  |  |
| 11-13 years | 25 (44.6) | 31 (55.4) | 19.05 | 0.000<sup>a</sup> |
| 14-16 years | 148 (65.2) | 79 (34.8) |  |  |
| 17-19 years | 127 (76) | 40 (24) |  |  |
| Gender |  |  |  |  |
| Male | 195 (64.3) | 110 (35.7) | 2.49 | 0.132<sup>b</sup> |
| Female | 102 (71.8) | 40 (28.2) |  |  |
| Education |  |  |  |  |
| J.S.C/8<sup>th</sup> grade | 27 (42.2) | 37 (57.8) | 50.03 | 0.000<sup>a</sup> |
|  | Present (%)<br>(n = 300) | Absent (%)<br>(n = 150) |  |  |
| S.S.C/O-levels | 77 (55) | 63 (45) |  |  |
| H.S.C/A-levels | 83(72.2) | 32 (27.8) |  |  |
| Under graduation | 113 (86.3) | 18 (13.7) |  |  |
| Monthly family income (BDT)<sup>c</sup> |  |  |  |  |
| Below 20000 BDT | 31 (36.9) | 53 (63.1) | 42.82 | 0.000<sup>a</sup> |
| 20000-40000 BDT | 131 (70.4) | 55 (29.6) |  |  |
| 41000-60000 BDT | 95 (77.2) | 28 (22.8) |  |  |
| >60000 BDT | 43 (75.4) | 14 (24.6) |  |  |
| Body Mass Index |  |  |  |  |
| Underweight | 67 (58.3) | 48 (41.7) | 9.10 | 0.011<sup>a</sup> |
| Normal | 213 (70.8) | 88 (29.2) |  |  |
| Overweight | 13 (50) | 13 (50) |  |  |
| <sup>a</sup>Pearson Chi-square; <sup>b</sup>Fisher’s exact correction test used for having 20% or more expected frequencies less than 5; <sup>c</sup>1 US dollar = 121.85 BDT (in May 2025) |  |  |  |  |

Table 5 revealed a highly significant association between LBP and both playing experience (p<0.001) and the duration of practice per week (p<0.001). A clear dose-response relationship is visible for both factors. LBP prevalence rises steadily with more years of playing, from 56.7% for those with 1-3 years of experience to 82.1% for those with 7-10 years. Similarly, LBP prevalence increases with more intense weekly practice, from 47.1% for those training less than 10 hours to 81.7% for those training more than 20 hours.

**Table 5.**
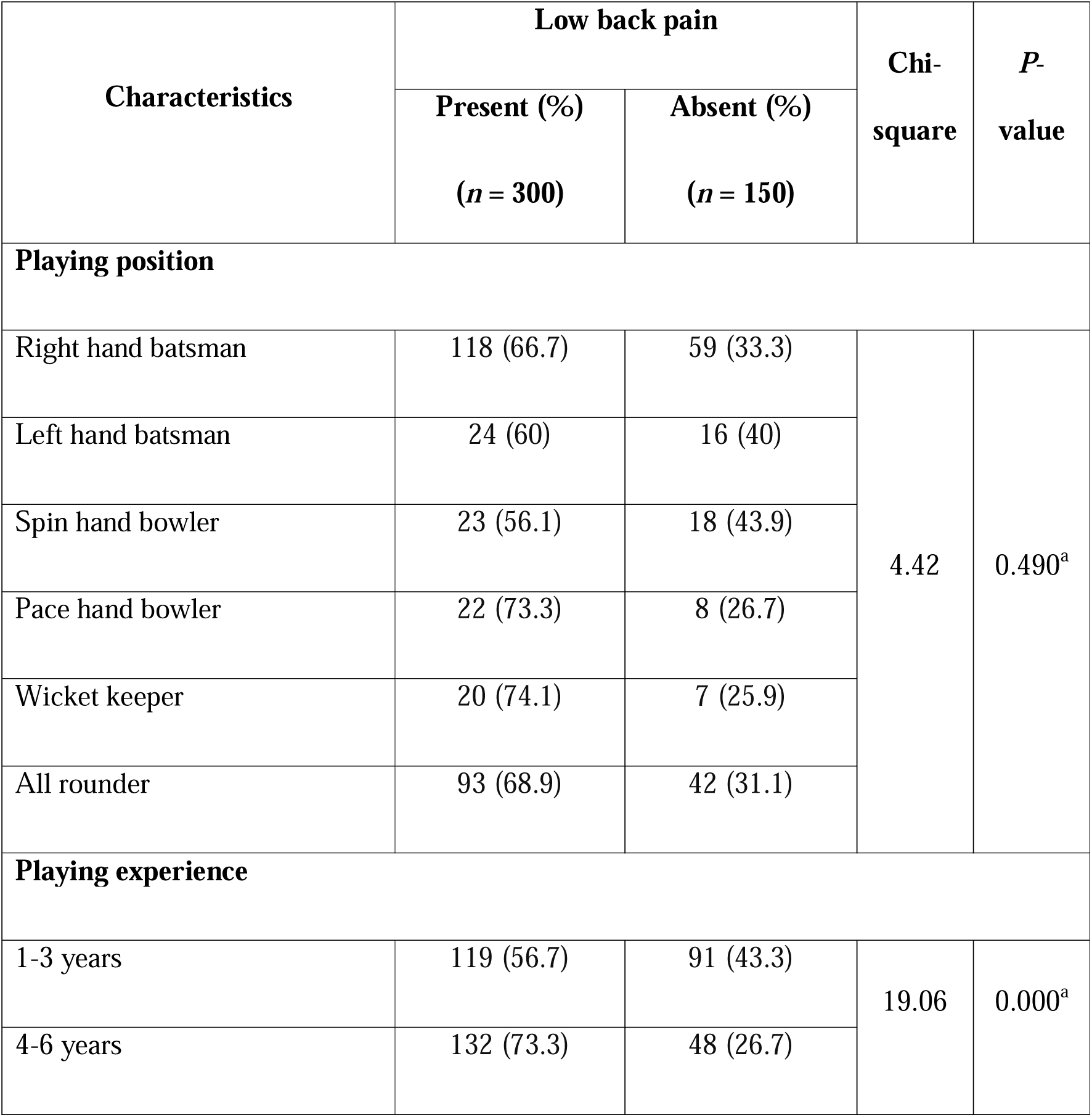
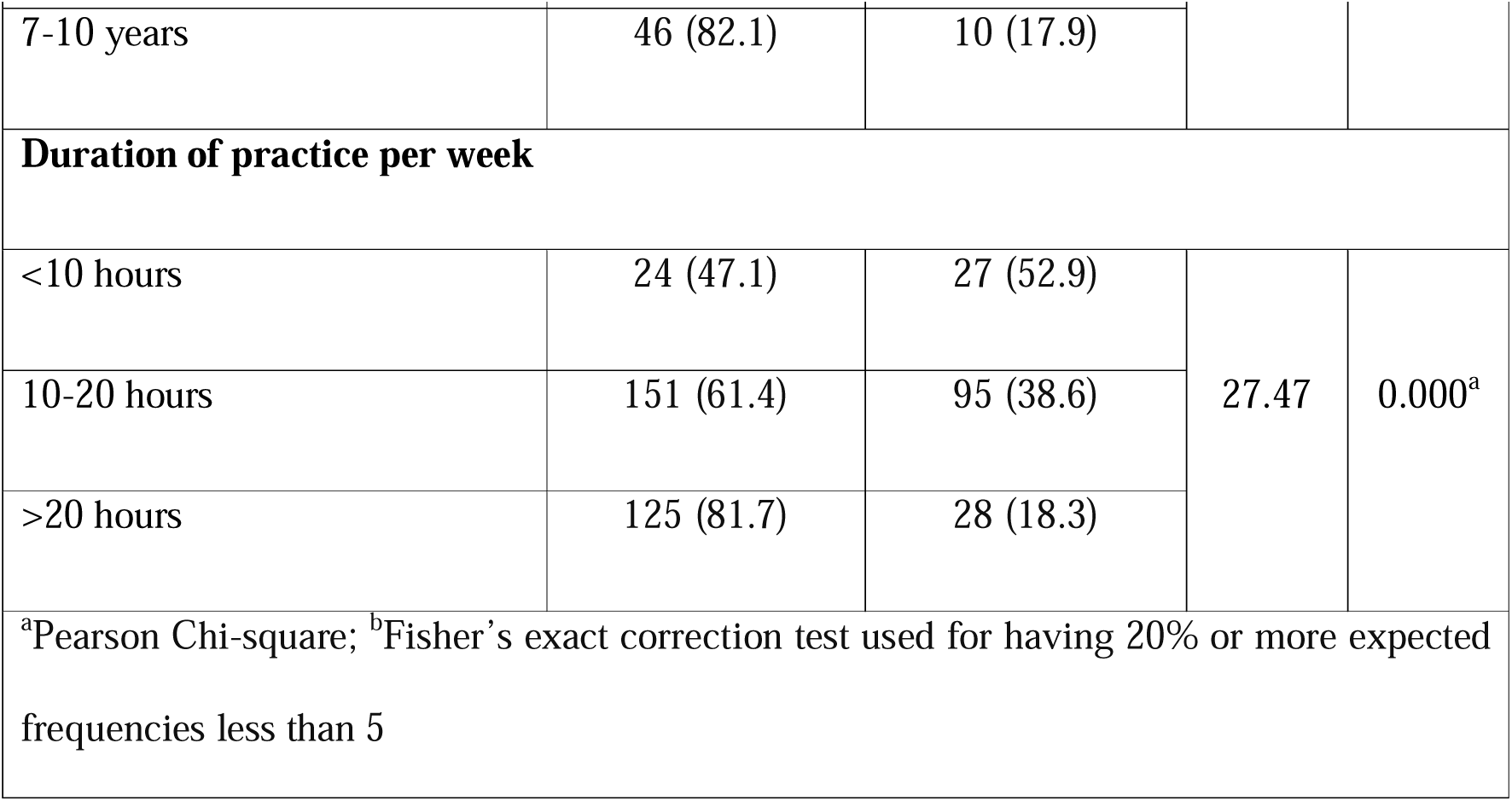
Distribution of games related characteristics of the participants (n=450)

Table 6 shows of both the frequency of warming up (p<0.001) and its duration (p<0.001) were significantly associated with LBP. The players who always warm up had a much lower prevalence of LBP (58.7%) than those who did so “Often” or “Sometimes” (96.3-97%). For duration, the group warming up for 10-15 minutes reported the highest rate of LBP (73.7%). Likewise, the frequency of cooling down after activity was highly significant (p<0.001), with those who “Always” cool down showing a markedly lower LBP prevalence (56.1%) compared to less consistent players. There is also a significant association between the duration of cool-down and low back pain (LBP) (p<0.001). The lowest prevalence of LBP (30.3%) is observed in the group of players who cool down for 10-15 minutes. In contrast, the prevalence of LBP is considerably higher for those with shorter cool-down periods of less than 10 minutes (61.4%) and is also elevated for those with longer cool-downs of more than 15 minutes (51.0%).

**Table 6.** Distribution of preventive measure related characteristics of the participants (n=450)

| Characteristics | Low back pain |  | Chi-square | P-value |
| --- | --- | --- | --- | --- |
|  | Present (%) | Absent (%) |  |  |
|  | (n = 300) | (n = 150) |  |  |
| Warm-up before sports activity |  |  |  |  |
| Always | 209 (58.7) | 147 (41.3) | 48.58 | 0.000<sup>a</sup> |
| Often | 26 (96.3) | 1 (3.7) |  |  |
| Sometimes | 65 (97) | 2 (3) |  |  |
| Duration of warm-up |  |  |  |  |
| <10 minutes | 77 (52.7) | 69 (47.3) | 19.08 | 0.000<sup>a</sup> |
| 10-15 minutes | 210 (73.7) | 75 (26.3) |  |  |
| >15 minutes | 13 (68.4) | 6 (31.6) |  |  |
| Cool-down after sports activity |  |  |  |  |
| Always | 169 (56.1) | 132 (43.9) | 45.41 | 0.000<sup>a</sup> |
| Often | 16 (84.2) | 3 (15.8) |  |  |
| Sometimes | 115 (88.5) | 15 (11.5) |  |  |
| Duration of cool-down |  |  |  |  |
| <10 minutes | 32 (88.9) | 4 (11.1) | 23.38 | 0.000<sup>a</sup> |
| 10-15 minutes | 219 (69.7) | 95 (30.3) |  |  |
|  | Present (%)<br>(*n* = 300) | Absent (%)<br>(*n* = 150) |  |  |
| >15 minutes | 49 (49.0) | 51 (51.0) |  |  |
| <sup>a</sup>Pearson Chi-square; <sup>b</sup>Fisher's exact correction test used for having 20% or more expected frequencies less than 5 |  |  |  |  |

### Logistic regression to predict risk factors of LBP

Table 7 presented the logistic regression analysis and this table revealed several key risk factors 236 for low back pain (LBP) among adolescent cricketers. In the regression model, it was revealed that inconsistent preventive habits were the most powerful predictor. Specifically, cricketers who sometimes (OR = 14.07, 95% CI: 2.78-71.13, p = 0.001) or often (OR = 12.48, 95% CI: 1.29-120.22, p = 0.029) warmed up had substantially higher odds of experiencing LBP compared to those who always did. Furthermore, the participants who cooled down sometimes nearly tripled the odds of LBP (AOR = 2.86, 95% CI: 1.34-6.09, p = 0.006). A previous history of LBP also emerged as a major risk factor, increasing the odds of a current episode nearly sevenfold (AOR = 6.92, 95% CI: 3.98-12.02, p = 0.010). Other significant independent predictors included female gender (AOR = 2.52, 95% CI: 1.25-5.07, p = 0.010), higher educational attainment as the undergraduate level (AOR = 5.38, 95% CI: 1.83-15.76, p = 0.002), and higher family income brackets.

**Table 7.**
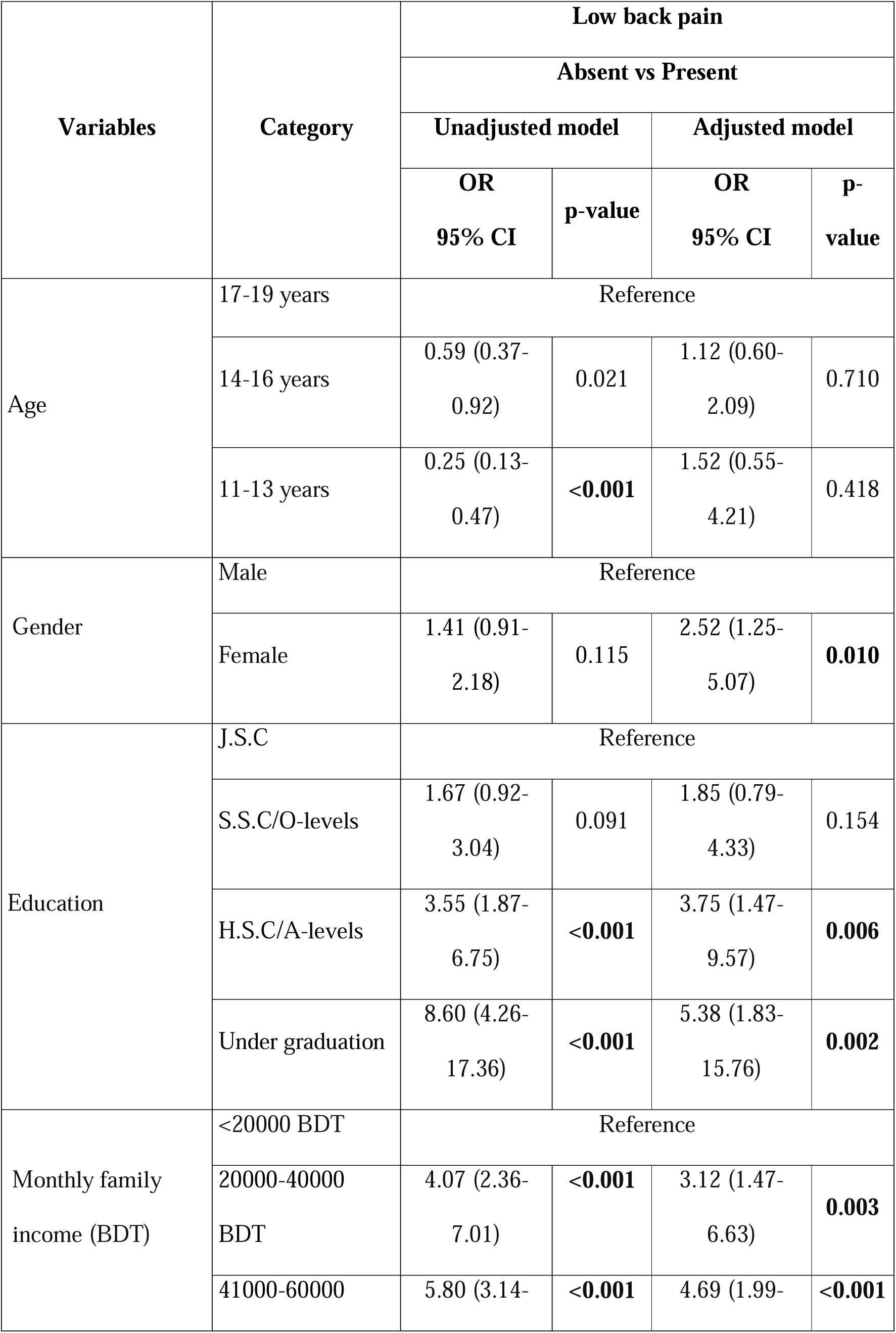
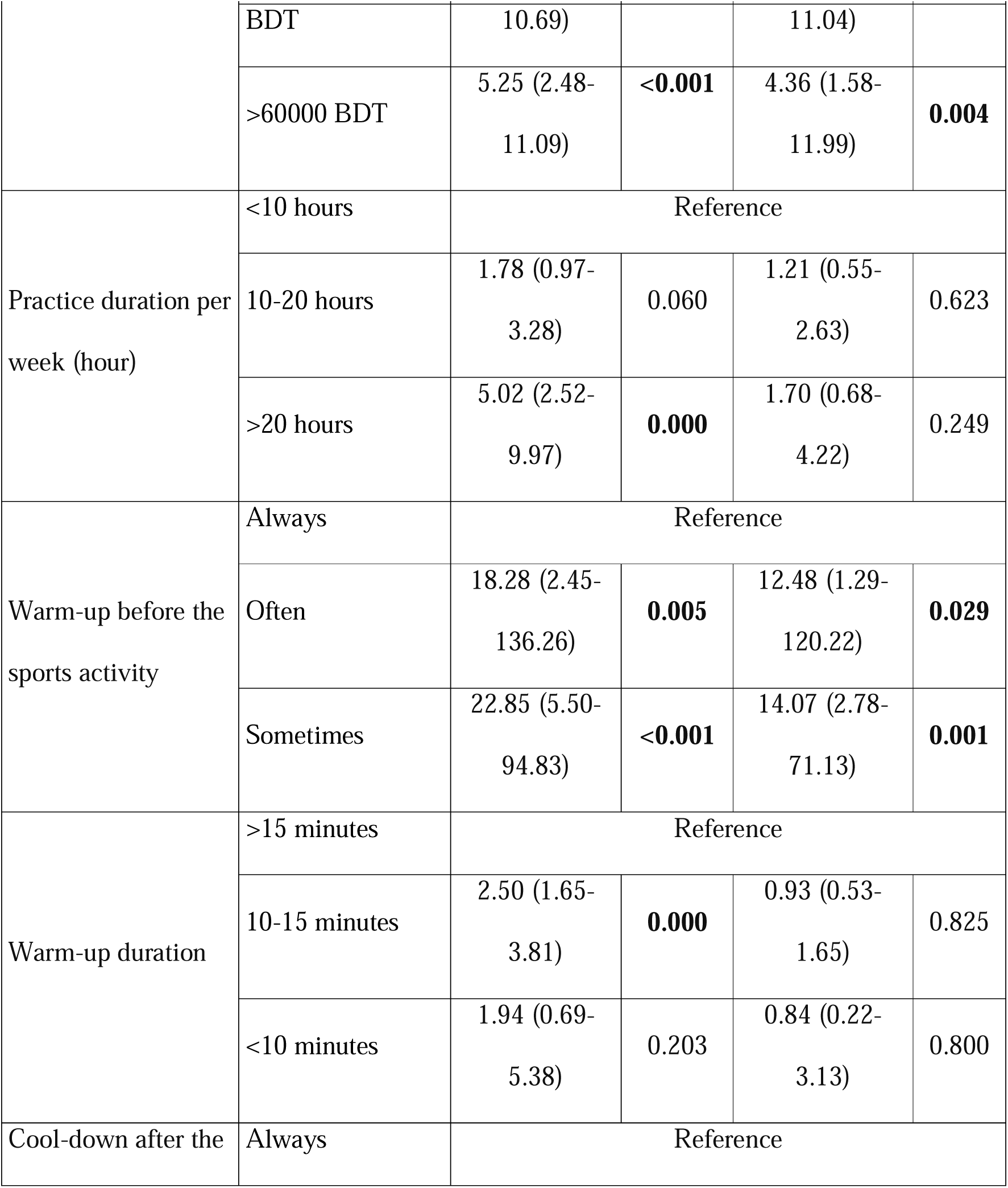
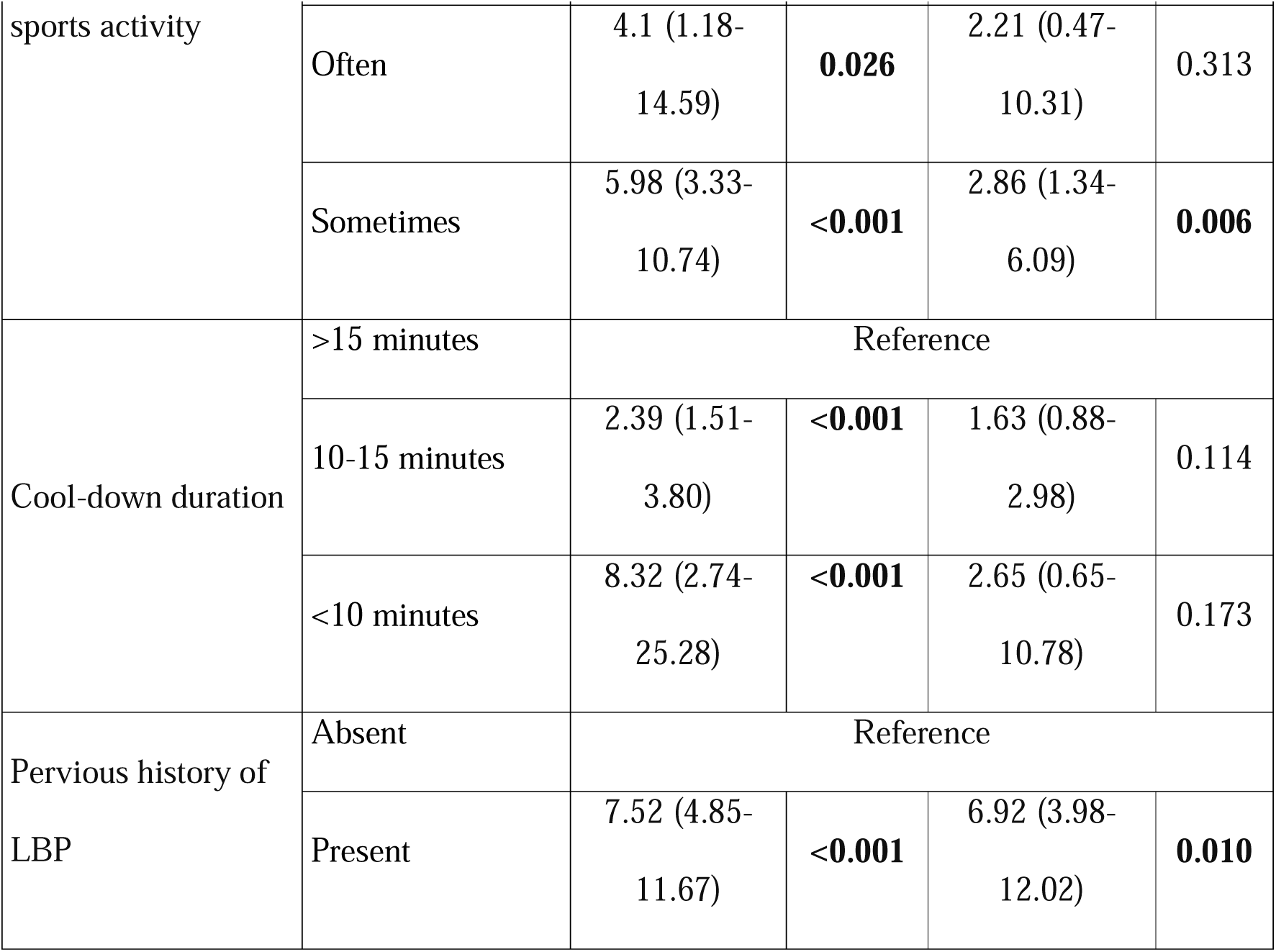
Logistic regression for sociodemographic and play related risk factors of low back pain among adolescent cricketers (n=450)

| Variables | Category | Low back pain |  |  |  |
| --- | --- | --- | --- | --- | --- |
|  |  | Absent vs Present |  |  |  |
|  |  | Unadjusted model |  | Adjusted model |  |
|  |  | OR<br>95% CI | p-value | OR<br>95% CI | p-value |
| Age | 17-19 years | Reference |  |  |  |
|  | 14-16 years | 0.59 (0.37-0.92) | 0.021 | 1.12 (0.60-2.09) | 0.710 |
|  | 11-13 years | 0.25 (0.13-0.47) | <0.001 | 1.52 (0.55-4.21) | 0.418 |
| Gender | Male | Reference |  |  |  |
|  | Female | 1.41 (0.91-2.18) | 0.115 | 2.52 (1.25-5.07) | 0.010 |
| Education | J.S.C | Reference |  |  |  |
|  | S.S.C/O-levels | 1.67 (0.92-3.04) | 0.091 | 1.85 (0.79-4.33) | 0.154 |
|  | H.S.C/A-levels | 3.55 (1.87-6.75) | <0.001 | 3.75 (1.47-9.57) | 0.006 |
|  | Under graduation | 8.60 (4.26-17.36) | <0.001 | 5.38 (1.83-15.76) | 0.002 |
| Monthly family income (BDT) | <20000 BDT | Reference |  |  |  |
|  | 20000-40000 BDT | 4.07 (2.36-7.01) | <0.001 | 3.12 (1.47-6.63) | 0.003 |
|  | 41000-60000 | 5.80 (3.14- | <0.001 | 4.69 (1.99- | <0.001 |
|  |  | Absent vs Present |  |  |  |
|  |  | Unadjusted model |  | Adjusted model |  |
|  |  | OR<br>95% CI | p-value | OR<br>95% CI | p-value |
|  | BDT | 10.69) |  | 11.04) |  |
|  | >60000 BDT | 5.25 (2.48-11.09) | <**0.001** | 4.36 (1.58-11.99) | **0.004** |
| Practice duration per week (hour) | <10 hours | Reference |  |  |  |
|  | 10-20 hours | 1.78 (0.97-3.28) | 0.060 | 1.21 (0.55-2.63) | 0.623 |
|  | >20 hours | 5.02 (2.52-9.97) | **0.000** | 1.70 (0.68-4.22) | 0.249 |
| Warm-up before the sports activity | Always | Reference |  |  |  |
|  | Often | 18.28 (2.45-136.26) | **0.005** | 12.48 (1.29-120.22) | **0.029** |
|  | Sometimes | 22.85 (5.50-94.83) | <**0.001** | 14.07 (2.78-71.13) | **0.001** |
| Warm-up duration | >15 minutes | Reference |  |  |  |
|  | 10-15 minutes | 2.50 (1.65-3.81) | **0.000** | 0.93 (0.53-1.65) | 0.825 |
|  | <10 minutes | 1.94 (0.69-5.38) | 0.203 | 0.84 (0.22-3.13) | 0.800 |
| Cool-down after the | Always | Reference |  |  |  |
|  |  | Absent vs Present |  |  |  |
|  |  | Unadjusted model |  | Adjusted model |  |
|  |  | OR<br>95% CI | p-value | OR<br>95% CI | p-value |
| sports activity | Often | 4.1 (1.18-14.59) | **0.026** | 2.21 (0.47-10.31) | 0.313 |
|  | Sometimes | 5.98 (3.33-10.74) | **<0.001** | 2.86 (1.34-6.09) | **0.006** |
| Cool-down duration | >15 minutes | Reference |  |  |  |
|  | 10-15 minutes | 2.39 (1.51-3.80) | **<0.001** | 1.63 (0.88-2.98) | 0.114 |
|  | <10 minutes | 8.32 (2.74-25.28) | **<0.001** | 2.65 (0.65-10.78) | 0.173 |
| Pervious history of LBP | Absent | Reference |  |  |  |
|  | Present | 7.52 (4.85-11.67) | **<0.001** | 6.92 (3.98-12.02) | **0.010** |

## Discussion

This research offers valuable findings regarding risk factors of low back pain (LBP) in adolescent cricket players of Dhaka City, applying machine learning to predictive modeling. The results highlighted the multi-factorial cause of LBP, stressing interaction between sociodemographic, game-related, and prevention-related factors.

Female gender was found to be an independent risk factor (AOR = 2.52, p = 0.010), aligning with general epidemiological trends where females often report higher rates of LBP, potentially due to anatomical, hormonal, or biomechanical differences. A large French occupational health study found female gender associated with increased risk of musculoskeletal disorder-related work unfitness (OR = 2.09) [18].

In this study, education level showed a strong and significant association with LBP (P<0.001). The participants having higher educational levels (Undergraduate) showing a greater prevalence of LBP (86.3%) compared to those with lower educational attainment. But a Brazilian study of 1,455 adolescents found that those who did not attend physical education classes had 2.26 times higher odds of experiencing LBP compared to participants [19].

In this study it was found that players who practiced > 20 hours/week had the highest prevalence of LBP (81.7%), while those who practiced <10 hours/week had the lowest prevalence (47.1%). Similarly, in a research, adolescent basketball players practicing over 6.6 hours/week exhibited 43.8% LBP prevalence [20] and youth weightlifters training long-term weightlifting developed lumbar disc degeneration within 4 years, progressing to herniation in 33% by year 5 [21].

A strong association was found between warming up before sports activity and LBP (P<0.001), with those who always warm up having a lower prevalence of LBP (56.6%). Similarly, a meta-analysis of Warm-up Intervention Programs (WIPs) found a 36% reduction in sports injuries when accounting for hours of risk exposure [22].

A strong association was observed between cool-down after sports activity and LBP (P<0.001), with those who always perform a cool-down having a lower prevalence of LBP (56.1%). Similarly, a randomized crossover study compared aqua- and land-based cool-down exercises found both have similar recovery effects on muscle soreness and performance-based parameters [23].

Previous history of LBP was a consistent and strong predictor of current LBP, with participants who had a prior history of LBP showing higher odds of presenting LBP (OR = 7.52). A comprehensive systematic review investigating the risk of LBP recurrence found that “a history of previous episodes of LBP prior to the most recent episode was the only factor that consistently predicted recurrence of LBP” [24]. Another systematic review also discovered intrinsic factors related to LBP in cricket fast bowlers to include previous injury as a factor known to influence load tolerance and LBP injury [25].

The application of machine learning here is a useful sports medicine innovation. Methods of traditional risk factor analysis cannot account for such complex, non-linear interdependencies amongst a large number of variables such as sociodemographic, game and prevention-of-injury factors and LBP factors.

## Conclusion

The aim of this study was to perform a risk assessment and classification of LBP among adolescent cricketers in Dhaka City, Bangladesh. This was an effort to fill an existing knowledge gap through ML techniques to derive key risk factors of LBP in this population. A wide range of variables were considered, including sociodemographic, game-related, preventive measures, and LBP-related variables, to evaluate model performance. A regression model was proposed to predict LBP risk factors, while a SVM model was identified as the most suitable ML classifier for the classification of LBP levels. LBP was classified into three levels, i.e., no pain, mild pain, and moderate pain, excluding severe pain category due to a limited number of severe cases. Despite the overall satisfactory performance, results show that identifying Class 2 (moderate pain) remains challenging. This may be due to overlapping clinical features between mild and moderate pain that limit separability. The findings provide significant insight to health professionals, coaches, and sport organizations in designing effective interventions to prevent and treat LBP in young players. Further research is required to evaluate the model across more diverse populations and incorporate additional discriminative features to better assess its generalizability for clinical applications.

## Supporting information

Supplement: Informed Consent form

Supplement: Questionnaire

## Data Availability

All data produced in the present study are available upon reasonable request to the authors

## Limitation

Non probability sampling, self-reported data

## Availability of data and materials

All supporting data are available on request from the authors.

## Acknowledgement

The authors would like to thank coaches and players of BKSP, Lt. Sheikh Jamal cricket Academy, Kola Bagan Krira chakra, Khelaghar Cricket Academy, and City club for all assistance and cooperation throughout the study.

## Supporting information

S1 Appendix. Informed consent form. The written consent form was administered to the participants.

S2 Appendix. Questionnaire. The included questions comprised the questionnaire administered to the participants.

S3 Appendix. Data. This file contains research data.

## Author Contribution

**Conceptualization:** Marzana Afrooj Ria, Tasrima Trisha Ratna, Shudeshna Chakraborttye Purba, Rubal Kar, Dr. Mohoshina Karim, Md Osman Ali, Erfat Jaren Chaity, Shahadath Hossen, Joynal Abedin Imran, Shahriar Hasan

**Data Curation:** Erfat Jaren Chaity, Shahriar Hasan

**Formal Analysis:** Tasrima Trisha Ratna, Rubal Kar

**Investigation:** Marzana Afrooj Ria, Shahriar Hasan

**Methodology:** Marzana Afrooj Ria, Md Osman Ali, Shudeshna Chakraborttye Purba

**Project Administration:** Dr. Mohoshina Karim, Joynal Abedin Imran

**Resources:** Rubal Kar, Erfat Jaren Chaity, Dr. Mohoshina Karim

**Software:** Md Osman Ali, Shahadath Hossen, Joynal Abedin Imran

**Supervision:** Marzana Afrooj Ria, Shahriar Hasan, Shudeshna Chakraborttye Purba

**Validation:** Tasrima Trisha Ratna, Shudeshna Chakraborttye Purba

**Visualization:** Md Osman Ali, Shahadath Hossen, Rubal Kar

**Writing – Original Draft:** Marzana Afrooj Ria, Tasrima Trisha Ratna

**Writing – Review & Editing:** Dr. Mohoshina Karim, Shahadath Hossen, Md Osman Ali

## Funding

No specific funding was allotted for this study.

## Notes

### Competing Interest Statement

The authors have declared no competing interest.

### Funding Statement

This study did not receive any funding

### Author Declarations

the Institutional Review Board (IRB) of the National Institute of Traumatology and Orthopaedic Rehabilitation (NITOR/PT/93/lRB/2024/05) gave ethical approval for this work.

### Summary of Updates

Methodology section has been updated to clarify; Figure 1,2 revised.

## Reference

1. Gera A, Pereira PC, Eapen C. Low Back Pain in Adolescent Athletes; Evaluation and Rehabilitation. In: Journal of Exercise Science and Physiotherapy. 2015;76. 10.18376//2015/v11i2/67706

2. Wall J, McGowan E, Meehan W, Wilson F. “Back pain is part of sport … I’m just gonna have to live with it”: Exploring the lived experience of sport-related low back pain in adolescent athletes. Phys Ther Sport. 2023;62:71–8. 10.1016/j.ptsp.2023.05.005

3. Kikuchi R, Hirano T, Watanabe K, Sano A, Sato T, Ito T, et al. Gender differences in the prevalence of low back pain associated with sports activities in children and adolescents: a six-year annual survey of a birth cohort in Niigata City, Japan. BMC Musculoskelet Disord. 2019;20(1):327. 10.1186/s12891-019-2707-9

4. Vij N, Naron I, Tolson H, Rezayev A, Kaye AD, Viswanath O, et al. Back pain in adolescent athletes: a narrative review. Orthop Rev. 2022;14(3). 10.52965/001c.37097

5. Orchard J, Saw R, Kountouris A, Redrup D, Farhart P, Sims K. Management of lumbar bone stress injury in cricket fast bowlers and other athletes. South Afr J Sports Med. 2023;35(1). 10.17159/2078-516X/2023/v35i1a15172

6. Senington B, Lee RY, Williams JM. Biomechanical risk factors of lower back pain in cricket fast bowlers using inertial measurement units: a prospective and retrospective investigation. BMJ Open Sport — Exerc Med. 2020 Aug 13;6(1):e000818. 10.1136/bmjsem-2020-000818

7. Desouza C, Jani C, Patil V. Non-Isthmic Spondylolysis Imaging Features: A Case Report. J Orthop Case Rep. 2022;12(02):42–4. 10.13107/jocr.2022.v12.i02.2658

8. Seas A, Zachem TJ, Valan B, Goertz C, Nischal S, Chen SF, et al. Machine learning in the diagnosis, management, and care of patients with low back pain: a scoping review of the literature and future directions. Spine J. 2025;25(1):18–31. 10.1016/j.spinee.2024.09.010

9. Abdollahi M, Ashouri S, Abedi M, Azadeh-Fard N, Parnianpour M, Khalaf K, et al. Using a Motion Sensor to Categorize Nonspecific Low Back Pain Patients: A Machine Learning Approach. Sensors. 2020;20(12):3600. 10.3390/s20123600

10. Bayne H, Elliott B, Campbell A, Alderson J. Lumbar load in adolescent fast bowlers: A prospective injury study. J Sci Med Sport. 2016;19(2):117–22. 10.1016/j.jsams.2015.02.011

11. Hossen S, Ali MO, Rashid MA, Higa H. Development of proning pose classification system for selfcare of COVID-19 patients. Prog Eng Sci. 2025;2(2):100064. 10.1016/j.pes.2025.100064

12. Panda NR. A Review on Logistic Regression in Medical Research. Natl J Community Med. 2022; 13(4):265–70. 10.55489/njcm.134202222

13. Xing W, Bei Y. Medical Health Big Data Classification Based on KNN Classification Algorithm. IEEE Access. 2020; 8:28808–19, 2020, 10.1109/ACCESS.2019.2955754

14. Apao NJ, Feliscuzo LS, Romana CL, Tagaro J. Multiclass classification using random forest algorithm to prognosticate the level of activity of patients with stroke. Int. J. Sci. Technol. Res. 2020; 9(4):1233–40. https://api.semanticscholar.org/CorpusID:216654256

15. Abe S. Support Vector Machines for Pattern Classification. London: Springer London; 2010. 10.1007/978-1-84996-098-4

16. Mohsen S, Elkaseer A, Scholz SG. Human Activity Recognition Using K-Nearest Neighbor Machine Learning Algorithm. In: Smart Innovation, Systems and Technologies. Singapore: Springer Singapore; 2022;304–13. 10.1007/978-981-16-6128-0_29

17. Xu L, Yang W, Cao Y, Li Q. Human activity recognition based on random forests. In: 2017 13th international conference on natural computation, fuzzy systems and knowledge discovery (ICNC-FSKD). IEEE; 2017;548–53. 10.1109/FSKD.2017.8393329

18. Pucci C, Dell’Omo M, Lesage F-X, Murgia N, Annesi-Maesano I, O-028 MUSCULOSKELETAL DISORDERS AS A WORK-RELATED RISK FACTOR OF UNFITNESS FOR WORK: A CROSS-SECTIONAL STUDY OF 1,327,540 WORKERS IN OCCITANIA, FRANCE. Occup. Med. 2024; 74: 0–0. 10.1093/occmed/kqae023.0487

19. Bergmann GG, Pinheiro E dos S, Mello JB, Graup S. Associação entre diferentes domínios da atividade física e a dor lombar inespecífica em adolescentes. Motricidade. 2020;16(3):245–54. 10.6063/motricidade.18141

20. Arend M, Toomsalu L, Kaasik P. High prevalence of ankle, knee and low back problems in highly trained adolescent basketball players at the beginning of their competitive season. Pap Anthropol. 2024;33(1):72–84. 10.12697/poa.2024.33.1.04

21. Yoshimizu R, Nakase J, Yoshioka K, Shimozaki K, Asai K, Kimura M, et al. Incidence and temporal changes in lumbar degeneration and low back pain in child and adolescent weightlifters: A prospective 5-year cohort study. PLoS ONE. 2022;17(6):e0270046. 10.1371/journal.pone.0270046

22. Ding L, Luo J, Smith DM, Mackey M, Fu H, Davis M, et al. Effectiveness of Warm-Up Intervention Programs to Prevent Sports Injuries among Children and Adolescents: A Systematic Review and Meta-Analysis. Int J Environ Res Public Health. 2022;19(10):6336. 10.3390/ijerph19106336

23. Chin ECY, Chung-Nam Lai S, Tsang SF, Ho-Ngai Chung S, Wong YL, Sran N, et al. Comparing the Effects of Aqua- and Land-Based Active Cooldown Exercises on Muscle Soreness and Sport Performance: A Randomized Crossover Study. Int J Sports Physiol Perform. 2024;19(12):1381–90. 10.1123/ijspp.2024-0020

24. da Silva T, Mills K, Brown BT, Herbert RD, Maher CG, Hancock MJ. Risk of Recurrence of Low Back Pain: A Systematic Review. J Orthop Sports Phys Ther. 2017;47(5):305–13. 10.2519/jospt.2017.7415

25. Farhart P, Beakley D, Diwan A, Duffield R, Rodriguez EP, Chamoli U, et al. Intrinsic variables associated with low back pain and lumbar spine injury in fast bowlers in cricket: a systematic review. BMC Sports Sci Med Rehabil. 2023;15(1):114. 10.1186/s13102-023-00732-1

