## Supplement: Questionnaire for "Machine Learning-Based Pattern Recognition of Risk Factors for Low Back Pain among Adolescent Cricket Players in Dhaka City"

**Participant ID No**:

**Name of the respondent:** **Date:**

**Institute/Club:**

**Present address of the respondent**:

**Section 1: Socio-Demographic Variable (s)**

| **Sl. No.** | **QUESTION** | **RESPONSE** | **Code No.** |
| --- | --- | --- | --- |
| 1 | Age Of the respondent? | _________years |  |
| 2 | Sex of the respondent? | ▭ Male  ▭ Female | 0  1 |
| 3 | Education | ▭ J.S.C/8^th^ grade  ▭ S.S.C/O-levels  ▭ H.S.C/A-levels  ▭ Under graduation | 0  1  2  3 |
| 4 | Monthly Family income? | _______________ BDT |  |
| 5 | Height | _______ Cm |  |
| 6 | Weight | _______ Kg |  |
| 7 | BMI (Body Mass index) | _______ Kg/m^2^ |  |

**Section 2: Play related variable (s)**

| **Sl. No.** | **QUESTION** | **RESPONSE** | **Code No.** |
| --- | --- | --- | --- |
| 8 | Which position do you play? | ▭ Right hand batsman  ▭ Left hand batsman  ▭ Spin hand bowler  ▭ Pace hand bowler  ▭ Wicket-keeper  ▭ All Rounder | 0  1  2  3  4  5 |
| 9 | Playing how many years have you been playing (sports experience)? | _________years |  |
| 10 | How long did you practice per week? | _________Hours |  |

**Section 3: Preventive measures related variable (s)**

| **Sl. No.** | **QUESTION** | **RESPONSE** | **Code No.** |
| --- | --- | --- | --- |
| 11 | Did you take warm up before match or competition? | ▭ Yes, Always  ▭ Yes, Often  ▭ Yes, sometimes  ▭ Never | 0  1  2  3 |
| 12 | (If the 11 is yes, then answer 12)  How long did you do warm up before match or competition? | __________Minutes |  |
| 13 | Did you take cool down after match or competition? | ▭ Yes, Always  ▭ Yes, Often  ▭ Yes, sometimes  ▭ Never | 0  1  2  3 |
| 14 | (If the 13 is yes, then answer 14)  How long did you do cool down after match or competition? | __________Minutes |  |

**Section 4: Low back pain related Variable (s)**

| **Sl. No.** | **QUESTION** | **RESPONSE** | **Code No.** |
| --- | --- | --- | --- |
| 15 | Do you have low back pain? | ▭ Yes  ▭ No | 0  1 |
| 16 | The severity of pain?  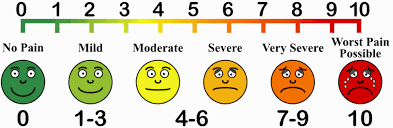 | ▭ No  ▭ Mild  ▭ Moderate  ▭ Severe | 0  1  2  3 |
| 17 | Previous episode of low back pain? | ▭ Yes  ▭ No | 0  1 |
